# Real Time Breast Histology Image Classification with a Mobile Phone

**DOI:** 10.1101/2022.09.15.21253543

**Authors:** Kenji Ikemura

**Affiliations:** Montefiore Medical Center

## Abstract

**Background:** Deep learning, specifically convolutional neural network, has made a breakthrough in the complex task of computer image recognition. In this study, depthwise separable convolutional neural network (DS-CNN), MobileNets v1, was used in classifying breast cancer histology images on the computer and then on a mobile/smart phone, in real time. This study propose that DS-CNN can be applied for histological image analysis and its network can be transferred to a commercially available mobile phone for real-time histological image analysis captured through the mobile phone camera.

**Method:** This study utilizes the DS-CNN on breast cancer histology images downloaded from publicly available repository: https://rdm.inesctec.pt/dataset/nis-2017-003. Training set images are augmented by rotation and mirroring the images. DS-CNN is trained to classify breast tissue images between 4 categories: i) normal, ii) benign, iii) carcinoma in-situ, and iv) invasive carcinoma. Finally, the trained DS-CNN is deployed on to a mobile phone to classify the images captured through the mobile phone camera in real-time. The output on the mobile phone screen is the real-time image from the camera and its probability of it being one of the 4 categories (from high to low confidence). Accuracy of DS-CNN is assessed, both on the computer and on mobile phone, by whether its prediction with highest confidence matches the true class in the test dataset. Secondary results of sensitivity and specificities were calculated.

**Results:** The trained DS-CNN accuracy on the computer reached as high as 86% in 4 class classification. On the mobile phone, accuracy reached 67% in 4 class classification and 78% in 2 class classification (normal or benign vs. in situ or invasive). Training time took less than 30 min on 1.4 GHz Intel Core i5 dual-core CPU. Latency for evaluation on mobile phone was less than 1 second. Demo video of real time histology image analysis with mobile phone can be found here: https://www.youtube.com/watch?v=qx2CdrSuazg

**Conclusion:** DS-CNN is a fast and efficient neural network architecture that can learn to distinguish histological images even with limited sample size and computational power. The architecture can be deployed onto a mobile phone and maintain relatively good accuracy through the phone camera. It is likely that the accuracy can be increased by expanding the dataset and with updated CNN that are optimized for mobile phones.

## I. Background

In the domain of medical imaging, convolutional neural network (CNN) has been successful applied in wide range of subspecialties from head to toe. In the field of pathology, CNN application has significantly expanded with advent of digital pathology. These applications include, but not limited to, classification of cancer type and counting mitosis.

However, some drawbacks of deep learning are that it requires large data samples, computational power, and time to reach high accuracy. Particularly for medical applications, it often requires thousands of images to train the computer and each image has a large file size. CNN is computationally demanding, and it often requires a GPU to do parallel processing, and yet it can still take days to complete the training.

This study demonstrates that depthwise separable convolutional neural network (DS-CNN), inspired by MobileNets (1), can classify breast histology images despite limited training sample size, computational power, and time. The study trained the network to classify between i) normal breast tissue, ii) benign, iii) in situ carcinoma and iv) invasive breast carcinoma. Finally, the trained network was transferred to a mobile phone to classify histological images through the mobile phone camera in real-time.

## II. Method

### Tools

The computer used was MacBook Air with 1.4 GHz Intel Core i5 dual-core CPU and the mobile phone was Samsung Galaxy S6 edge+ with an Exynos 7420 octa-core CPU (2.1GHz quad + 1.5GHz quad) and with 4 GB RAM, 16-megapixel camera (2988 × 5312 pixels).

### Dataset

The training and test datasets were downloaded from publicly available dataset provided by Araújo et al (2): https://rdm.inesctec.pt/dataset/nis-2017-003. This dataset is composed of high-resolution (2040 × 1536 pixels), uncompressed Tagged Image File Format (TIF) format, and annotated Hematoxylin and Eosin (H&E) stain images. All images are digitized with the same acquisition conditions, with a magnification of 200x and pixel size of 0.42*μm* x 0.42*μm*. Training dataset is composed of four classes of breast tissue: 55 normal, 69 benign, 63 in situ, and 62 invasive breast cancers. Representative images are shown in figure 1. Total of 249 unique training images were prepared. For test set, unique images were prepared with 5 images from each class of breast tissue, total of 20 images, which were denoted as “Initial” test dataset. An additional test set composed of unique 16 images (4 images from each category), were provided with images of increased ambiguity, which were denoted as “extended” test dataset. Total of 36 images were prepared for test set. The labeling was performed by two pathologists, who only provided a diagnosis from the image contents, without specifying the area of interest for the classification. Cases of disagreement between specialists were discarded.

**Figure 1:**
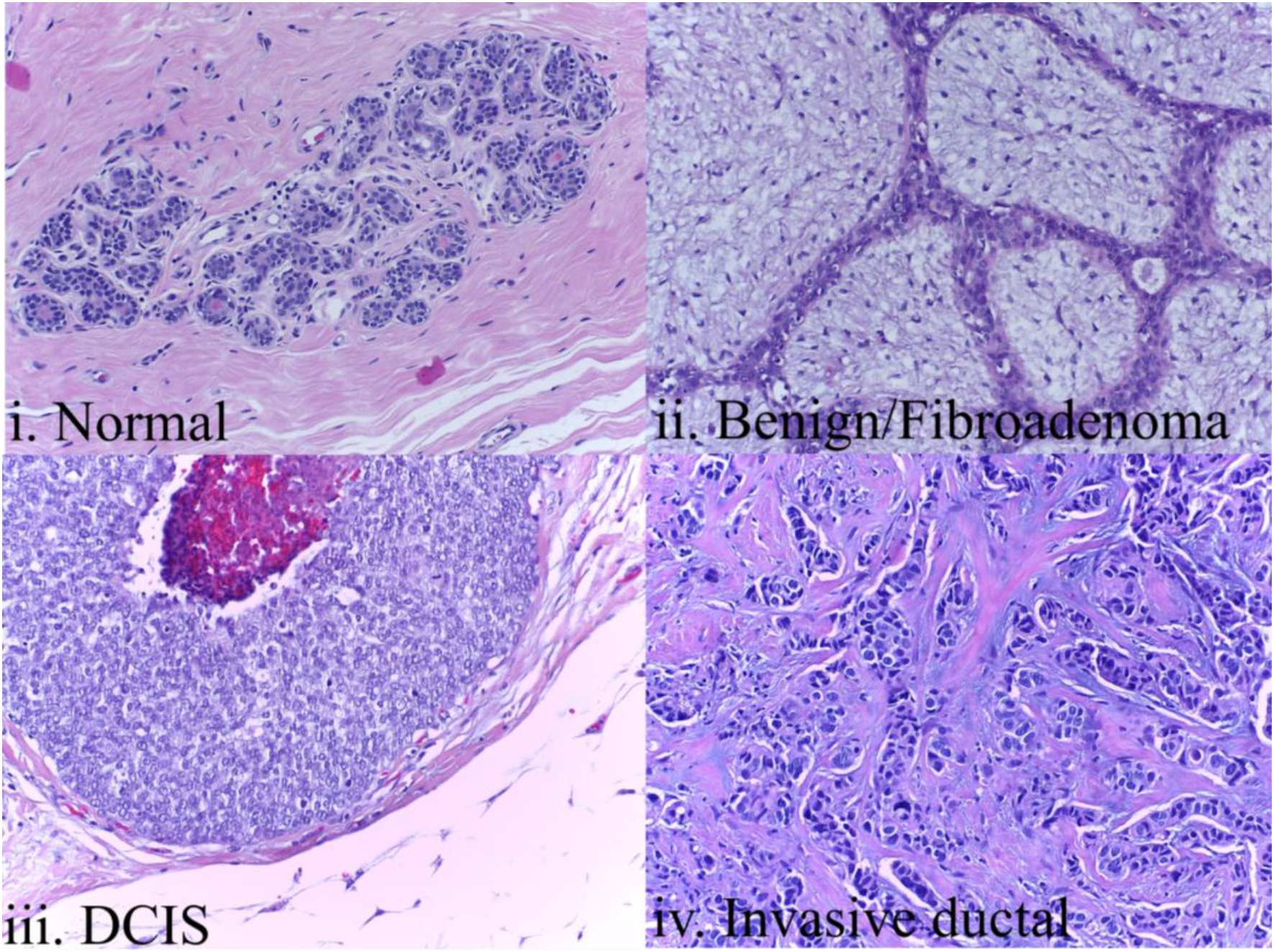
Representative images of the 4 breast tissue histology classes.

### Image augmentation

First, all images were converted from TIF format to JPEG format with minimal loss. This reduced the size of an image from 9MB to about 5 MB each. Due to low number of sample size, training dataset was augmented by adding images of *k* · *π/4* rotations, with k = {0, 1, 2, 3}, and horizontal reflections. This resulted in 249 × 4 × 2 = 1992 training samples.

### MobileNets v1 and Deployment of model to Mobile Phone

For this study, we used a DS-CNN designed by Google named MobileNets v1 (1). MobileNets After training the neural network on the computer, we deployed the final layer of trained network on to the Samsung Galaxy S6 Edge+ smart phone. The test set images were presented on iPad. The phone was set right above the iPad, about 19cm of space in between. This allowed the entire image displayed on iPad to show on the mobile phone (Figure 2).

**Figure 2:**
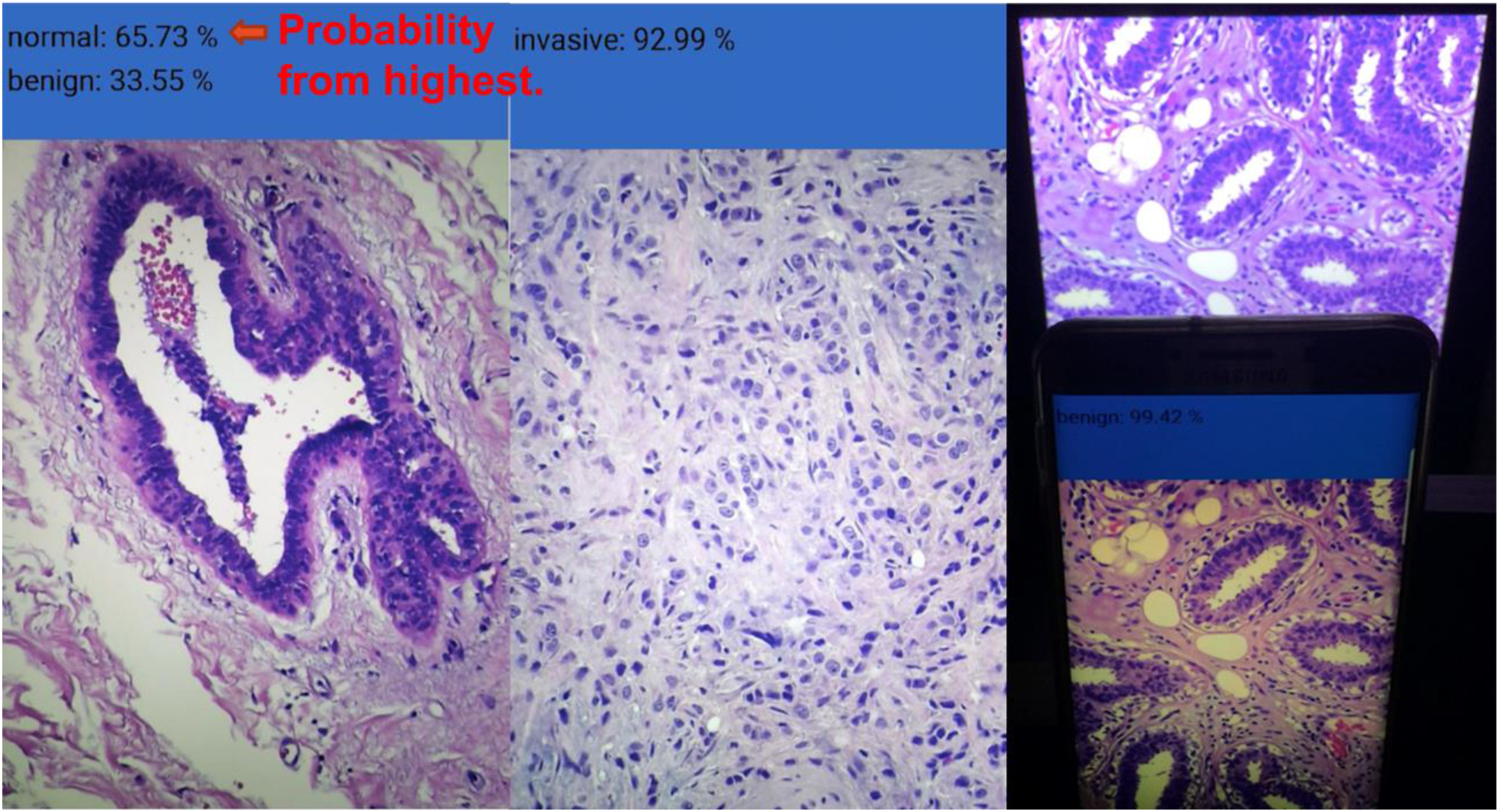
The left two images are representation of what the mobile phone screen shows when presented with breast tissue. The image farthest right is the setup of how breast tissue image on ipad was presented to the mobile phone.

### Results evaluation

The performance of our method is evaluated in terms of accuracy, sensitivity, and specificity. This evaluation is performed image-wise for the initial and extended sets. A quaternary and binary classification was tested. For binary classification, normal and benign breast tissue was combined into one category as non-carcinoma, while in situ and invasive breast cancer were combined into one category as carcinoma. Accuracy of DS-CNN is assessed, both on the computer and on mobile phone, by whether its prediction with highest confidence matches the true class in the test dataset.

## III. Results

The time it took to train the network was approximately 30 min. The resulting DS-CNN model size was 17MB. This model was deployed on to the mobile phone. Final training set accuracy was 100% and final cross-validation accuracy was 97.8%. The cross entropy of training and validation test set was 0.018 and 0.06, respectively (Figure 3). The overall accuracy, sensitivity, and specificity on the test set for computer and mobile phone are presented in Table 1, 2 and 3, respectively. Demo video of real time histology image analysis with mobile phone can be found here: https://www.youtube.com/watch?v=qx2CdrSuazg

**Table 1:** Accuracy

|  | 4 Classes |  |  | 2 Classes |  |  |
| --- | --- | --- | --- | --- | --- | --- |
|  | Initial Test set | Extended Test set | Overall | Initial Test set | Extended Test set | Overall |
| Computer | 90 | 81.3 | 86.1 | 100 | 93.8 | 97.2 |
| Mobile phone | 70 | 62.5 | 67 | 85 | 68.8 | 78 |

**Table 2:**
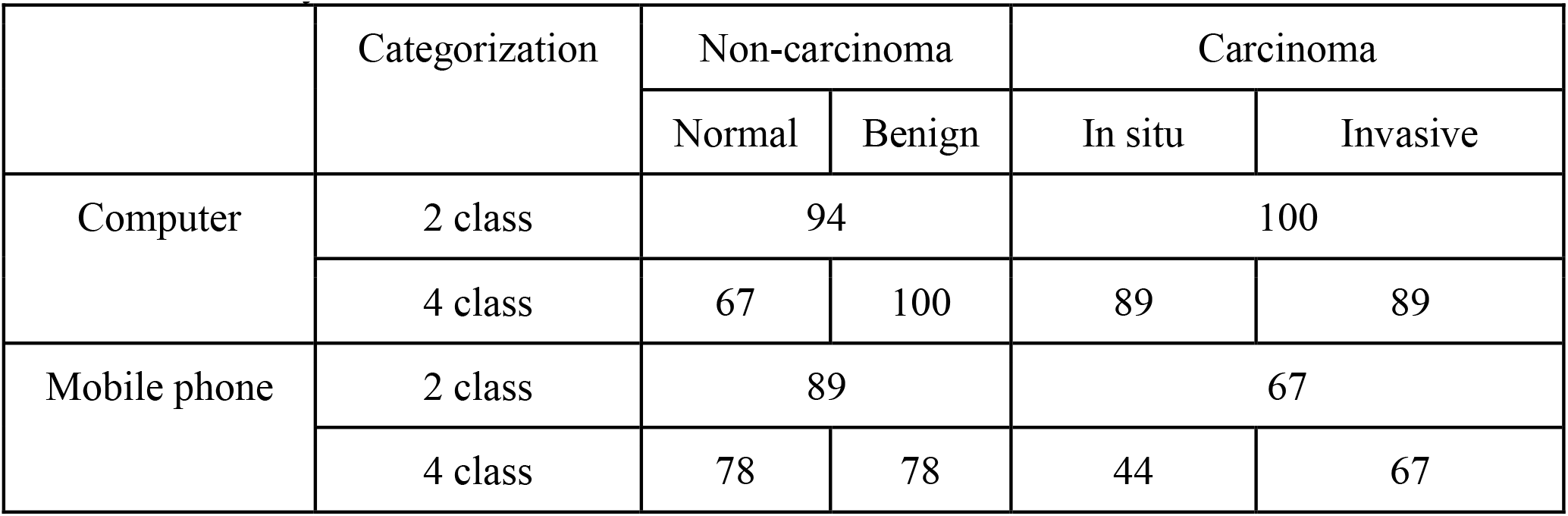
Sensitivity

**Table 3:**
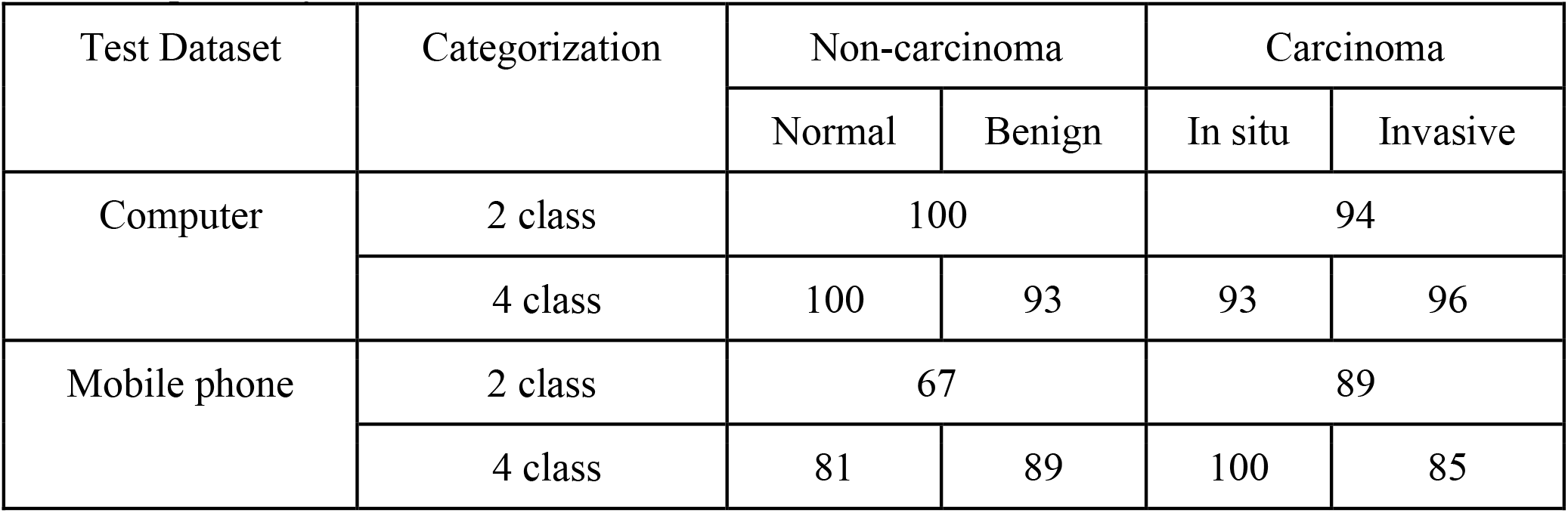
Specificity

**Figure 3:**
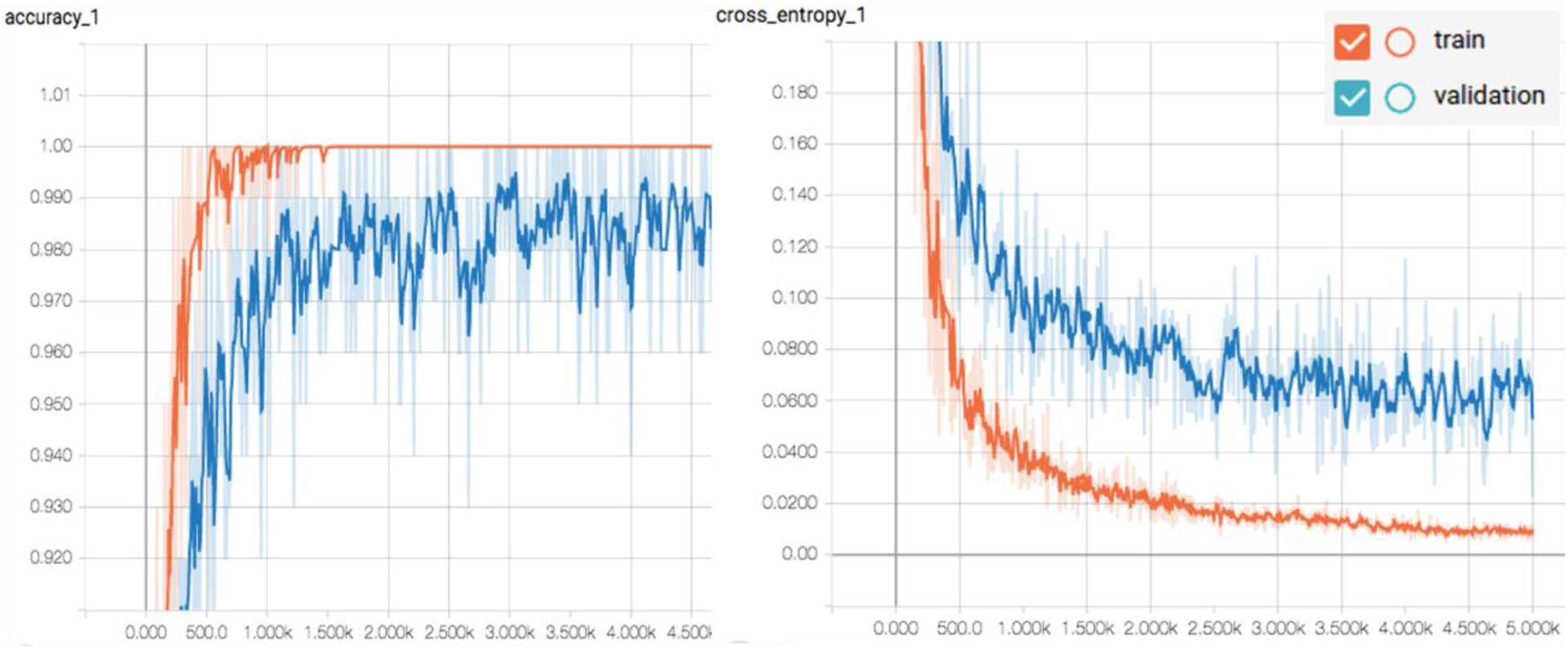
The cross entropy of training and validation test set was 0.018 and 0.06, respectively.

## IV. Discussion

This study trained DS-CNN to classify breast histology images and attained high accuracy, sensitivity, and specificity. The simplicity of DS-CNN made it easy and fast to run the experiment even on a 1.4 GHz CPU. The last layer of the trained DS-CNN was successfully deployed to a mobile phone, and it maintained decent levels of performance in real-time.

It is likely that there is some level of overfitting in the training due to limited sample size. Many regularization techniques such as dropout, drop connect, and weight decay, attempts to solve the problem of over-fitting by reducing the capacity of their respective models. However, these methods may reduce the capacity of the neural network model and simplify the complexity of the learning problem. Larger training sample size can overcome overfitting. This can be done by collecting more unique images or by image augmentation techniques such as patching, finer rotation, or by adding random noise into the images. Patching techniques, as seen in Araujo et al, can also allow the network to characterize subnuclear features such as nuclear membrane and chromatins (2). Nevertheless, despite limited sample size and without patching, our network was able to perform at high accuracy.

On the mobile phone, accuracy of 4 class accuracy classification was 67% and 2 class accuracy classification was 78%. There are still challenges that must overcome to increase accuracy on a mobile phone. Issues such as the camera quality, surrounding light and glare, and even small movement of the camera can influence the image captured for the CNN to analyze in real-time. In the future, standardized mobile phone settings may be necessary when using mobile phone cameras for medical diagnosis. Nevertheless, this study proves the concept that histological image analysis on a mobile phone via deep learning is possible.

Utilization of mobile phone for medical diagnosis may become useful in rural areas where there are less pathologists or where there are no local high-end computers. Because the main CNN training can be performed on to the computer and then transferred to the mobile phone, it does not require physicians to have high-end computers in their local clinic. The application may also have use in education. There are still many challenges ahead to realize a useful mobile application that can do histological image analysis, such as building a user interface. However, the study results indicate that histological image analysis via mobile phone camera is possible.

### Peak into CNN’s Blackbox

The difficulty of unraveling CNN’s decision making is inherent in its architecture. The computer is looking for low level features such as edges and curves, and then building up to more abstract concepts through series of non-linear functions.

In this study, all images were converted to 246 × 246 pixels, and because patching was not implemented, it is likely that the network is looking solely at tissue organization features of the entire image rather than submicroscopic features (eg, nuclear features). It is possible that the DS-CNN has trained itself to pick up features like what pathologist would use to distinguish these 4 categories. Picture labeled as normal tissue had many lobular structures that had rather distinct boarder between the lobules and surrounding stroma. Cluster of grapes like structure is unique to healthy breast tissue and would likely be easy for the network to catch through edge detection.

Benign tissues, on the other hand, were more likely to be filled with fat and fibrous tissue. Ducts are compressed into slit-like space by the proliferative stroma and branching leaf like glands are apparent. In both, normal and benign tissues, the structures have distinct boarders with paler stroma flowing in a uniform direction compared to carcinoma that appears more disorganized.

For benign images, on the computer, all benign images in test set were diagnosed correctly from initial and extended test sets. There were 2 false positives that were diagnosed as benign when it was normal breast tissue. There was a normal tissue that was misdiagnosed as in-situ in a rather fatty tissue. Perhaps the computer perceived the fat lobules as a strong sign of normal than carcinoma. This was a case from extended test set that was difficult even for a trained pathologist to make a correct diagnosis just from a single image.

For in situ images, since cellular feature and subnuclear feature are unlikely to be detected, the network likely did not catch the pleomorphism and prominent nucleoli of cells. However, the smooth circular boarder from intact myepithelium may have trigged DS-CNN to call it as in-situ. In addition, there were many cribriform structures in training and test set that may have helped our network to associate this morphology with in-situ. Just for fun, we presented the trained network with an image of a sushi roll, and it classified it as in-situ (Figure 4). There were 2 false positive where normal breast tissue was called invasive. It is possible that the DS-CNN considered fibrous tissue patterns a strong feature of normal tissue. While fibrous pattern of carcinoma was that of desmoplasia.

**Figure 4:**
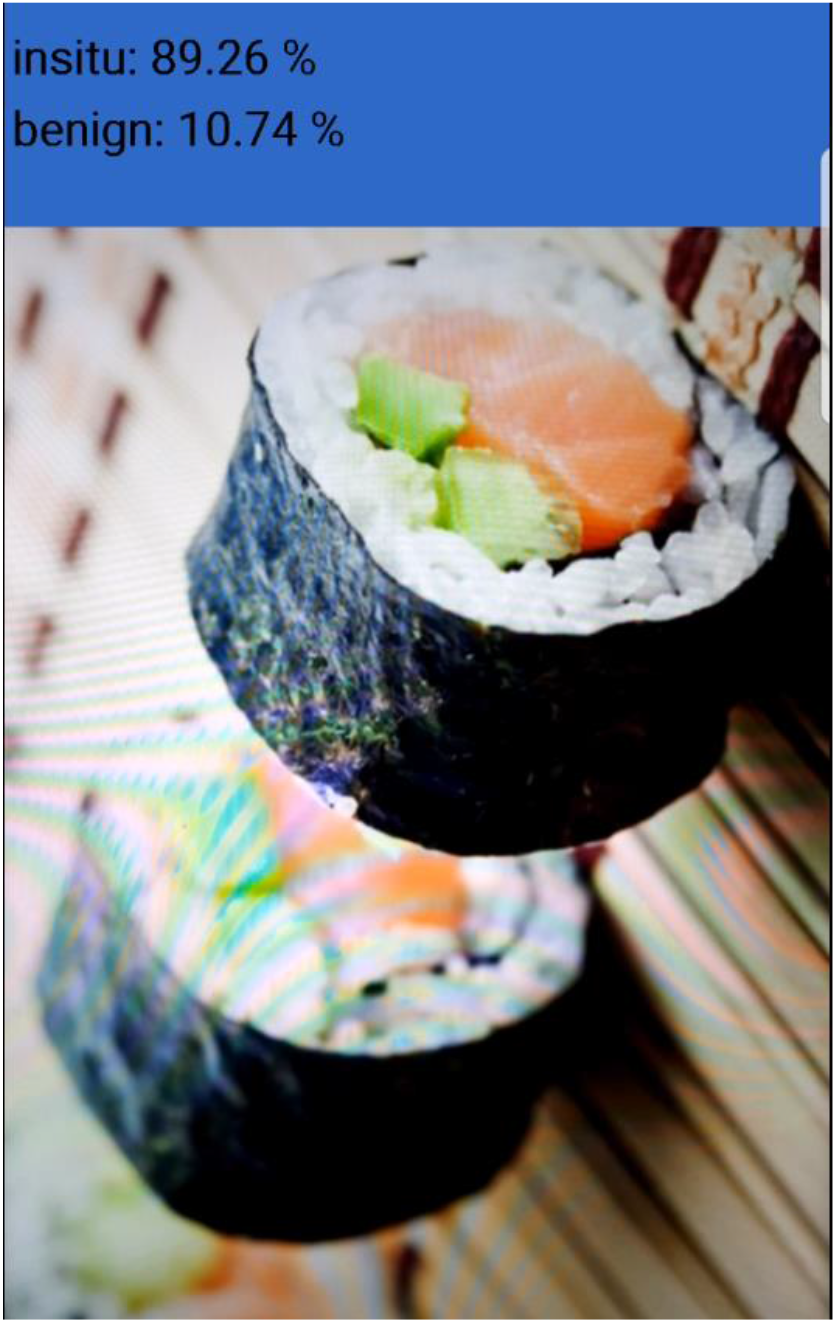
When sushi roll was presented to the mobile phone camera, the DS-CNN considered it as in-situ breast tissue.

The DS-CNN had an easier time identifying invasive carcinoma from its disorganized and dense scar tissues (desmoplastic reaction). Tubules are radiating outwards in a stellate fashion with irregular boarders. It is less likely for DS-CNN to have been able to recognize the heterogeneity of cells commonly seen in carcinomas. However, overall morphological features of invasive structures are quite eye-catching even to an untrained eye and perhaps to our network as well.

For invasive cancer images, all initial invasive images were diagnosed correctly, and one extended case was misdiagnosed as in situ. There was one false positive that was actually an in-situ case. Perhaps a focus of microinvasion was detected. However, it is a reasonable mistake between invasive versus in situ from these single shot images.

It was a nice surprise to find DS-CNN (MobileNets v1) able to perform at high accuracy despite the complex nature of histology images. Even a well-trained pathologist will have difficulty making definitive diagnosis just from looking at these single shot images. For example, some invasive images in test set had fat lobules that can look like normal or benign tissue, but DS-CNN correctly classified it as invasive carcinoma. It is likely that it is using other criteria than from what pathologist generally use to classify these images. If we increase the training sample size through patching and train with subnuclear information, the accuracy may further improve.

## V. Conclusion

DS-CNN has achieved high accuracy, sensitivity, and specificity in classifying breast tissue despite limited training data, computational power, and time. The final layer of the network was successfully deployed to a mobile/smart phone and maintained decent accuracy and sensitivity. In the future, with larger data set, GPU, and architecture optimization (e.g. number of layers, nodes, etc.), it will certainly be possible for computers to make more consistent and precise diagnosis, even on a smart phone.

## Data Availability

Data downloaded from publicly available repository.

https://rdm.inesctec.pt/dataset/nis-2017-003

